# The kinetics of humoral and cellular responses after the booster dose of COVID-19 vaccine in inflammatory arthritis patients

**DOI:** 10.1101/2022.12.28.22284008

**Authors:** Jakub Wroński, Bożena Jaszczyk, Leszek Roszkowski, Anna Felis-Giemza, Krzysztof Bonek, Anna Kornatka, Magdalena Plebańczyk, Tomasz Burakowski, Barbara Lisowska, Brygida Kwiatkowska, Włodzimierz Maśliński, Małgorzata Wisłowska, Magdalena Massalska, Ewa Kuca-Warnawin, Marzena Ciechomska

## Abstract

**Introduction:** Impaired immunogenicity of COVID-19 vaccinations in inflammatory arthritis (IA) patients results in diminished immunity. However, optimal booster vaccination regimens are still unknown, due to not unstudied kinetics of the immune response after booster vaccinations. Therefore, this study aimed to assess the kinetics of humoral and cellular responses in IA patients after the COVID-19 booster.

**Patients and Methods:** In 29 IA patients and 16 healthy controls (HC) humoral responses (level of IgG antibodies) and cellular responses (IFN-γ production) were assessed before (T0), after 4 weeks (T1), and after more than 6 months (T2) from the booster vaccination with BNT162b2.

**Results:** IA patients, but not HC, showed lower anti-S-IgG concentration and IGRA fold change at T2 compared to T1 (p=0.026 and p=0.031). Furthermore, in IA patients the level of cellular response at T2 returned to the pre-booster level (T0). All immunomodulatory drugs, except IL-6 and IL-17 inhibitors for the humoral and IL-17 inhibitors for the cellular response, impaired the immunogenicity of the booster dose at T2. However, none of the immunomodulatory drugs affected the kinetics of both humoral and cellular responses (measured as the difference between response rates at T1 and T2).

**Conclusion:** Our study showed impaired kinetics of both humoral and cellular responses after the booster dose of the COVID-19 vaccine in IA patients, which, in the case of cellular response, did not allow the vaccination effect to be maintained for more than 6 months. Repetitive vaccination with subsequent booster doses seems to be necessary for IA patients.

## Introduction

Previous studies have established impaired immunogenicity of COVID-19 vaccinations in inflammatory arthritis (IA) patients after the primary vaccination schedule. Lower humoral and cellular responses compared to healthy controls (HC) are caused mainly due by the immunomodulatory treatment IA patients received (1,2). The abrogated immunological response in IA patients after the primary vaccination schedule (3–5) caused the necessity of booster vaccinations. Current studies proved that, though COVID-19 booster vaccinations are effective in IA patients(6–8), also the immunogenicity of the booster dose is impaired in this group of patients(9– 12). In our previous study, we assessed humoral and cellular responses before the booster dose (more than 6 months from the primary vaccination schedule; T0) and 4 weeks after the booster dose (T1) in 49 IA patients and 47 HC vaccinated at the COVID-19 vaccination unit in a rheumatology center (13). Our study showed a boost of humoral and cellular responses in IA patients, but lower than in HC. The humoral response after the booster dose was only impaired by biological and targeted synthetic disease-modifying antirheumatic drugs (bDMARDs and tsDMARDs), but the cellular response was decreased after all immunomodulatory drugs except IL-17 inhibitors and sulfasalazine. However, optimal booster vaccination regimens are still unknown, due to a lack of data regarding the kinetics of immunological response after the booster dose. Therefore, we continued the observation of part of the previously described cohort, and this study aimed to assess the kinetics of humoral and cellular responses in IA patients after the booster dose of the COVID-19 vaccine.

## Patients and methods

The study protocol was approved by the hospital bioethics committee (KBT-3/2/2021). All participants signed informed consent for inclusion in the study. In 29 IA patients and 16 HC, we assessed humoral and cellular immunity after more than 6 months from the booster dose (respectively median 201 and 199 days after; T2). The level of IgG antibodies against the SARS-CoV-2 Spike (S1) antigen was measured with anti-SARS-CoV-2 QuantiVac ELISA (Euroimmun, Lübeck, Germany) and the IFN-γ production with interferon-gamma release assay (IGRA) Quan-T-Cell SARS-CoV-2 (Euroimmun, Lübeck, Germany). To detect COVID-19 infection after the booster dose we also measured antibodies against SARS-CoV-2 nucleocapsid (N) with SARS-CoV N ELISA Kit (TestLine Clinical Diagnostics, Brno, Czech Republic). The compliance of the data with the normal distribution was assessed using the Shapiro–Wilk test. The significance of the observed differences between the two groups was assessed using the Student’s T test for variables with a normal distribution, the Mann–Whitney U test for variables without a normal distribution, and categorical variables the Fisher’s exact test. To assess the kinetics the Friedman test was used, with post hoc analysis with the Dunn’s test. The significance of the results after adjusting for confounding factors was checked by linear regression. Statistical analysis was performed using Statistica 13.3 software (StatSoft Polska, Cracow, Poland) and figures were created using GraphPad Prism 6 software (GraphPad Software, San Diego, CA, US).

## Results

Patient characteristics are presented in Table 1. There were no significant differences between the groups, apart from the older age of the IA group (p=0.002). In both groups, a similar percentage of subjects had COVID-19 after the booster dose, based on the presence of antibodies against SARS-CoV-2 nucleocapsid.

**Table 1.** Patient characteristics. bDMARDs – biological disease-modifying antirheumatic drugs, GCs – glucocorticoids, IL-6i – IL-6 inhibitors, IL-17i – IL-17 inhibitors, JAKi – JAK inhibitors, MTX – methotrexate, N/A – not applicable, nrSpA – non-radiographic spondylarthritis, ns – nonsignificant, SSA – sulfasalazine, TNFi – TNF inhibitors, tsDMARDs – targeted synthetic disease-modifying antirheumatic drugs.

|  | Inflammatory arthritis<br>(n=29) | Healthy control<br>(n=16) | difference |
| --- | --- | --- | --- |
| Age (mean ± SD) | 54.5 ± 14.2 | 41.8 ± 9 | p=0.002 |
| Sex – female (n, %) | 16 (55.2%) | 10 (62.5%) | ns |
| BMI (mean ± SD) | 27.2 ± 4.3 | 27.7 ± 5.6 | ns |
| Smoking (n, %) |  |  |  |
| - current | 3 (10.3%) | 1 (6.3%) | ns |
| - past | 8 (27.6%) | 1 (6.3%) | ns |
| Vaccine booster dose (n, %) |  |  |  |
| - Pfizer | 29 (100%) | 16 (100%) | ns |
| Heterologous Booster Vaccine (n, %) | 6 (20.7%) | 3 (18.8%) | ns |
| Days after booster vaccination (median, min, max) |  |  |  |
| - first point (T1) | 31 (22, 52) | 31 (27, 77) | ns |
| - second point (T2) | 201 (167, 290) | 199 (175, 234) | ns |
| COVID-19 infection after booster vaccination (n, %) | 5 (17.2%) | 3 (18.8%) | ns |
| Disease type (n, %) |  | - | N/A |
| - Rheumatoid arthritis | 16 (55.2%) |  |  |
| - Ankylosing spondylitis | 6 (20.7%) |  |  |
| - Psoriatic arthritis | 6 (20.7%) |  |  |
| - nrSpA | 1 (3.4%) |  |  |
| Immunomodulatory treatment (n, %) | 28 (96.6%) | - | N/A |
| GCs (n, %) | 11 (37.9%) | - | N/A |
| - dose (median, min, max) | 5 (2.5, 37.5) |  |  |
| cDMARDs (n, %) |  | - | N/A |
| - MTX | 17 (58.6%) |  |  |
| - SSA | 4 (13.8%) |  |  |
| bDMARDs and tsDMARDs (n, %) | 18 (62.1%) | - | N/A |
| - TNFi | 9 (31%) |  |  |
| - JAKi | 5 (17.2%) |  |  |
| - IL-6i | 2 (6.9%) |  |  |
| - IL-17i | 2 (6.9%) |  |  |
| bDMARDs and tsDMARDs used: |  |  |  |
| - in monotherapy | 11 (37.9%) |  |  |
| - with cDMARDs | 7 (24.1%) |  |  |
| Treatment suspension during booster vaccination (n, %) | 2 (6.9%) | - | N/A |

The kinetics of humoral response (anti-S-IgG concentration) after the COVID-19 booster vaccination is presented in Figure 1. At T2 levels of antibodies were significantly higher in both IA patients and HC than at T0 (p=0.001 and p=0.002 respectively), but lower in IA patients (median 406, min 10.7, max 5166.4) than in HC (median 1253.2, min. 331.4, max. 2561.9). The differences remained significant after adjustment for age (p=0.029). Additionally, in IA, but not in HC, a statistically significant decline (p=0.026) in the humoral response between T1 and T2 was observed, indicating a faster waning of humoral response in IA patients compared to HC.

**Figure 1.**
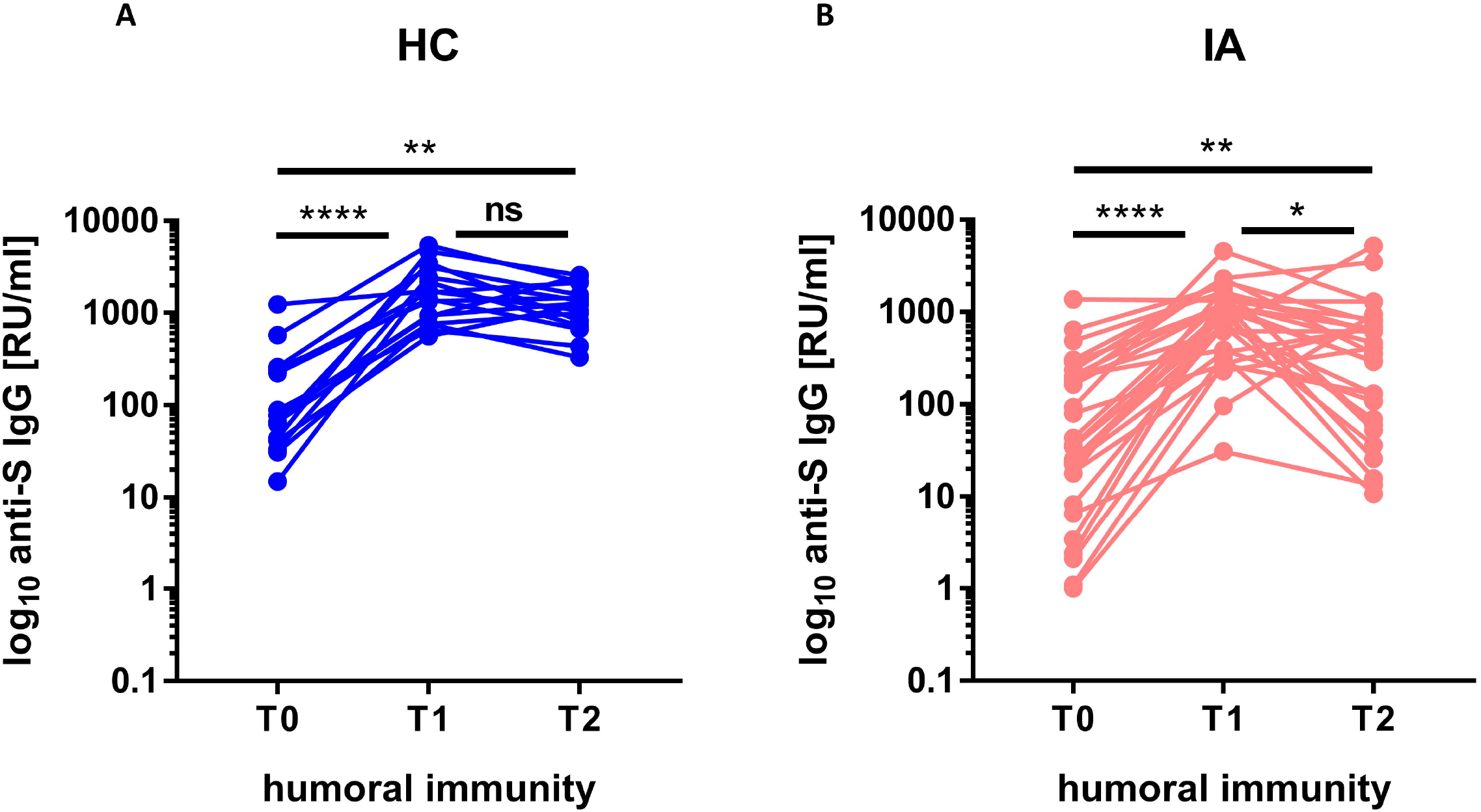
The level of anti-S IgG (humoral immunity) before (T0), after 4 weeks (T1), and after more than 6 months (T2) from the booster dose of the COVID-19 vaccine in healthy controls (HC; n=16) (A) compared to patients with inflammatory arthritis (IA; n=29) (B). In the group comparison, the Friedman test with the Dunn’s multiple comparison test was used. Dots represent individual values and p values were expressed as follows: 0.05>p>0.01 as*, 0.01>p> 0.001 as**; 0.001>p>0.0001as***, p<0.0001 as****, ns – nonsignificant.

The kinetics of cellular response (IGRA fold change) after the COVID-19 booster vaccination is presented in Figure 2. The cellular response at T2 was significantly higher than at T0 in HC (p=0.002), but not significantly different than at T1. However, in IA patients the level of cellular response at T2 was not only significantly lower in IA patients (median fold change 5, min. 0, max. 1763.8) than in HC (median 1236.7, min. 19.2, max. 30161, after adjustment for age p=0.004). It also significantly decreased from T1 to T2 (p=0.031), returning to the pre-booster level – there was no significant difference between the cellular response at T0 and T2 in IA patients. This fact indicates severely diminished kinetics of the cellular response in IA patients.

**Figure 2.**
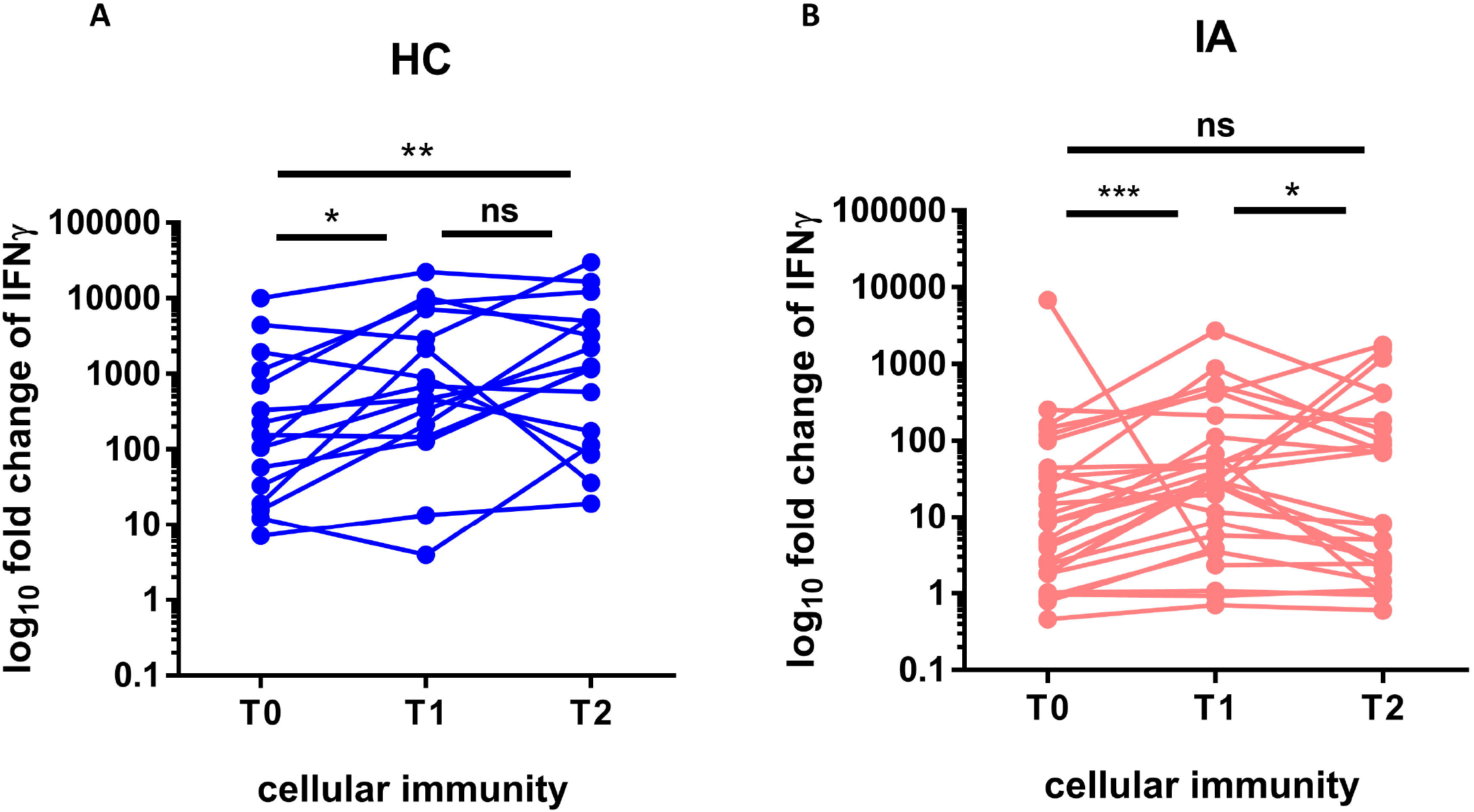
The fold change of IFNγ production stimulated by viral protein (cellular immunity) before (T0), after 4 weeks (T1), and after more than 6 months (T2) from the booster dose of the COVID-19 vaccine in healthy controls (HC; n=16) (A) compared to patients with inflammatory arthritis (IA; n=29) (B). In the group comparison, the Friedman test with the Dunn’s multiple comparison test was used. Dots represent individual values and p values were expressed as follows: 0.05>p>0.01 as*, 0.01>p> 0.001 as**; 0.001>p>0.0001as***, p<0.0001 as****, ns – nonsignificant.

The effect of each immunomodulatory drug on the immunogenicity of the COVID-19 vaccines at T2 is shown in Table 2. The analysis showed a significantly reduced humoral response in IA patients compared to HC after all drugs except IL-6 and IL-17 inhibitors and cellular response after all drugs except IL-17 inhibitors. All results of immunomodulatory drugs’ effect after the adjustment for age remained significant. There were no statistically significant differences in humoral and cellular responses between patients using bDMARDs and tsDMARDs in monotherapy vs in combination with conventional DMARDs. However, a significantly lower cellular (but not humoral) response was observed for IA patients using immunomodulatory drugs in combination with glucocorticoids (p<0.001). Interestingly, none of the immunomodulatory drugs affected the kinetics of both humoral and cellular responses after the booster dose (measured as the difference between response rates at T1 and T2).

**Table 2.** Effect of immunomodulating drugs on the immunogenicity of the COVID-19 vaccines more than 6 months from booster vaccination. bDMARDs – biological disease-modifying antirheumatic drugs, GCs – glucocorticoids, IL-6i – IL-6 inhibitors, IL-17i – IL-17 inhibitors, JAKi – JAK inhibitors, MTX – methotrexate, ns – nonsignificant, SSZ – sulfasalazine, TNFi – TNF inhibitors, tsDMARDs – targeted synthetic disease-modifying antirheumatic drugs.

| IgG antibodies, median RU/ml (min, max) |  |  |
| --- | --- | --- |
| Healthy control (HC, n=16) | 1253.2 (331.4, 2561.9) | The difference compared to HC |
| GCs (n=11) | 107.4 (10.7, 1273.4) | p<0.001 |
| MTX (n=17) | 406 (10.7, 5166.4) | p=0.007 |
| SSZ (n=4) | 452.8 (13.4, 778.1) | p=0.011 |
| bDMARDs and tsDMARDs (n=18) | 380.4 (10.7, 3476.4) | p<0.001 |
| - TNFi (n=9) | 293 (36.1, 1294.8) | p<0.001 |
| - JAKi (n=5) | 614.5 (107.4, 943.2) | p=0.008 |
| - IL-6i (n=2) | 351.9 (10.7, 693.1) | ns |
| - IL-17i (n=2) | 1915.6 (354.8, 3476.4) | ns |
| IGRA, median fold change (min, max) |  |  |
| Healthy control (HC, n=16) | 1236.7 (19.2, 30161) | The difference compared to HC |
| GCs (n=9) | 0.95 (0, 182.7) | p<0.001 |
| MTX (n=16) | 73.6 (0, 1763.8) | p=0.002 |
| SSZ (n=3) | 46.9 (0.9, 142.5) | p=0.016 |
| bDMARDs and tsDMARDs (n=18) | 6.3 (0, 1763.8) | p<0.001 |
| - TNFi (n=9) | 69.5 (0, 409.7) | p<0.001 |
| - JAKi (n=5) | 2.5 (0, 1186.7) | p=0.004 |
| - IL-6i (n=2) | 0.85 (0.6, 1.1) | p=0.013 |
| - IL-17i (n=2) | 1092.6 (421.4., 1763.8) | ns |

In our relatively small group of patients, we did not observe an effect of the level of humoral and cellular responses at T1 on the chances of new COVID-19 infection in the first 6 months after the booster vaccination.

## Discussion

Our study demonstrates impaired kinetics of both humoral and cellular responses after the booster dose of the COVID-19 vaccine in IA patients. The faster decline of the immune response compared to HC after the booster dose indicates the need for more frequent booster vaccinations in patients with IA than in HC. This is in line with the ACR recommendations, which, based on Centers for Disease Control and Prevention recommendations for non-immunocompetent people, currently recommends vaccinating patients with rheumatic diseases with 5 doses of the COVID-19 vaccine in total(14).

The waning of humoral immunity after the COVID-19 booster seems to be slower than after the primary vaccination regimen – the levels of responses were higher at T2 than at T0 and none of the IA patients in our study lost the humoral response after more than 6 months from booster vaccination (compared to 20.4% of IA patients after more than 6 months from the primary schedule as reported in our previous study). What’s more, although most of the immunomodulatory drugs used significantly reduced humoral and cellular response in IA patients compared to HC, both after primary and booster vaccinations, our study did not show their effect on the kinetics of the response. It may indicate that the kinetics impairment may result to a greater extent from dysfunction of the immune system in IA than from immunomodulatory drugs.

Our study, however, showed severe impairment in the kinetics of cell-mediated immunity after COVID-19 vaccination. After COVID-19 infection virus-specific T cell half-life is estimated to be of around 200 days(15). In our IA patients, the level of cellular response after 200 days from the booster dose returned to the pre-booster level (Figure 2). This indicates a severely impaired cellular response in patients with IA and should be taken into account when determining the optimal booster schedules.

The greatest advantage of our study is the assessment of the kinetics of both humoral and cellular responses in IA patients after the booster dose and the effect of individual immunomodulatory drugs on the kinetics. The obvious limitation of our study is a relatively small sample size and unmatched age of the groups (though we accounted for it in our analysis). Studies on larger groups of patients may allow the determination of the kinetics with a distinction between various types of IA and to detect (if existent but more subtle) the effect of waning immunity on the rate of COVID-19 infections. Finally, the exact levels of antibodies and cellular responses to protect against COVID-19 are still unknown.

Our study showed impaired kinetics of immune responses after the booster dose of the COVID-19 vaccine, which, in the case of cellular response, did not allow the vaccination effect to be maintained for more than 6 months. Periodic vaccination with subsequent booster doses seems to be necessary for IA patients. Further studies with longer follow-up periods should allow the determination of optimal intervals between vaccinations.

## Data Availability

Available upon reasonable request sent to the corresponding author.

## Author Contributions

Conceptualization, JW, BJ, and MC; methodology, JW, MM, MC and EK-W; validation, JW, MM, MC and EK-W; formal analysis, JW, MM, MC and EK-W; investigation, JW, BJ, LR, AF-G, AK, MP, TB, BL, MM, MC, EK-W; resources, BJ, AF-G, BK, WM, MW; data curation, JW, LR and BJ; writing – original draft preparation, JW, MM, MC and EK-W; writing – review and editing, BJ, LR, AF-G, KB, AK, MP, TB, BL, BK, WM, and MW; visualization, MM, MC and EK-W; supervision, BK, WM, and MW; project administration, JW and MC; funding acquisition, KB, BL, and BJ. All authors have approved the manuscript for submission.

## Data availability statement

Available upon reasonable request sent to the corresponding author.

## Ethics statement

The study protocol was approved by the National Institute of Geriatrics, Rheumatology and Rehabilitation bioethics committee (KBT-3/2/2021). All participants signed informed consent for inclusion in the study. The study was conducted according to the Declaration of Helsinki.

